# Type 1 Diabetes Mellitus in Tanzania: a systematic review of prevalence and incidence

**DOI:** 10.1101/2024.10.09.24315148

**Authors:** Lucy Elauteri Mrema, Anthony Mapunda, Pauline Sylvester, Willyhelmina Olomi, Nyanda Elias Ntinginya, Mary Mayige

**Affiliations:** National Institute for Medical Research- Mbeya Medical Research Centre, Tanzania; National Institute for Medical Research, Headquarters’- Dar es Salaam, Tanzania; Muhimbili University of Health and Allied Sciences, Tanzania

**Keywords:** Type 1 diabetes mellitus (T1DM), Incidence, Prevalence, Tanzania, Children

## Abstract

**Objective:** Type 1 diabetes mellitus (T1DM) is a chronic disease that affects children and adolescents globally. However, there is a paucity of data on the incidence and prevalence T1DM in Tanzania. This systematic review aimed to determine the prevalence and incidence of type 1 diabetes mellitus Tanzania through a comprehensive review of available literature.

**Methods:** A systematic literature search was conducted in six electronic databases (PubMed, Scopus, Web of Science, Embase, African Journals Online, and Tanzania Medical Journal) to identify studies reporting on the prevalence and incidence of T1DM in Tanzania. The search was conducted according to the Preferred Reporting Items for Systematic Reviews and Meta-Analyses (PRISMA) guidelines.

**Results:** A total of 63 studies were identified through the initial search, of which two studies met the inclusion criteria. Both studies were conducted in Dar es Salaam, Mwanza, Zanzibar, Kilimanjaro Tanzania, and reported on the incidence and prevalence of T1DM in children and adolescents. The studies were published in 1993 and 2019, respectively. The first study reported an annual incidence of T1DM of 1.5 per 100,000 population aged 0-19 years, while the second study reported an annual incidence ranging from 1.8 to 1.9 per 100,000 children and a prevalence ranging from 10.1 to 11.9 per 100,000 children.

**Conclusion:** The prevalence and incidence of T1DM in Tanzania are relatively low, based on the limited evidence available. More prevalence and incidence studies are needed to provide a better understanding of the burden of T1DM to inform diabetes management strategies in the country.

**Ethics:** Since this review has used previously published studies, consent of an ethics committee was not sought.

**PROSPERO registration number:** CRD42022369954.

## Introduction

Diabetes mellitus is a chronic metabolic disorder that has become a growing public health concern worldwide. Symptoms of diabetes include polyuria, increased thirst, visual disturbances, and weight loss (1). Severe presentations may include diabetic ketoacidosis and hyperosmolar states, potentially resulting in stupor and coma. Many symptoms may initially manifest mildly, yet harbour the propensity to inflict cumulative organ damage over time, predisposing individuals to irreversible complications such as vision impairment, limb amputation, stroke, and ultimately, fatality. Previously termed insulin-dependent diabetes, Type 1 diabetes may manifest at any age but predominantly afflicts children and young adults (2).

T1D specifically, has been relatively rare in sub-Saharan Africa, but recent studies have shown an increasing incidence and prevalence in children and adolescents in Tanzania (3,4). In Tanzania, the implementation of programs supporting children with Type 1 diabetes have contributed to a decline in morbidity and mortality rates despite the gradual increase in diabetes prevalence over recent decades(3). Despite this notable impact, diabetes often receives limited public health attention and insufficient research funding. One contributing factor to this oversight may stem from the substantial expenses associated with confirmatory diagnostic tests, particularly those assessing islet autoantibody levels, potentially leading to underestimated prevalence rates. Comprehensive information regarding the incidence and prevalence of Type 1 diabetes across different regions of Tanzania remains limited. Hence, there exists an urgency for accurate, evidence-based epidemiological data to guide planning and facilitate the formulation of tailored healthcare interventions. Therefore, this systematic review seeks to assess the available evidence concerning the incidence and prevalence of Type 1 diabetes in Tanzania.

## Methods

We conducted a systematic review to identify all relevant studies reporting on prevalence of T1D, or incidence of T1D, screening methods for type 1 diabetes (T1D), criteria used to diagnose T1D, in Tanzania. We excluded studies that focused on type 2 diabetes only, reviews on T1D, editorials, and molecular studies. There was no language or date limitations.

We searched six electronic databases (PubMed, Scopus, Web of Science, Embase, African Journals Online, and Tanzania Medical Journal) to identify eligible studies. The search was conducted in 1^st^ October 2023 to 15^th^ Dec 2023, without any time limits. We used the following search terms to identify relevant studies: type 1”[MeSH Terms] OR “Insulin Dependent Diabetes Mellitus 1”[Title/Abstract] OR “IDDM”[Title/Abstract] OR “Juvenile Onset Diabetes”[Title/Abstract] “Prevalence”[MeSH Terms] OR “Period Prevalence”[Title/Abstract] OR “Point Prevalence”[Title/Abstract] “Incidence”[MeSH Terms] OR “Incidence Proportion”[Title/Abstract] OR “Cumulative Incidence”[Title/Abstract] OR “Incidence Rate”[Title/Abstract] Tanzania [ MeSH Term (Appendix I, shows the PRISMA flow). We also searched for grey literature and manuscripts by manually searching reference lists and conference proceedings. Grey literature was included if it met the inclusion criteria, and its quality was assessed using the same the Joanna Briggs Institute (JBI) critical appraisal checklist.

Two authors independently screened the titles and abstracts of all identified studies for eligibility. Full texts of potentially relevant studies were then reviewed by the same two authors, and any discrepancies were resolved through discussion. We used the online screening tool Covidence http://www.covidence.org/) to manage the screening process, pre-populate inclusion and exclusion criteria, and remove duplicate records.

Data extraction was guided by JBI checklist for prevalence studies (Appendix II) and completed by two independent review authors. Any discrepancies were resolved through discussion or with a third reviewer. The following information was extracted: study identification (title, authors, year, journal), study design (type of study, setting, sampling method), participant characteristics (age, sex, ethnicity, inclusion/exclusion criteria), data collection methods, outcome measures (prevalence and/or incidence), and results.

The quality of studies was assessed using the JBI critical appraisal checklist for prevalence and incidence studies. The checklist consists of nine questions designed to assess the quality of studies reporting prevalence/incidence data, and studies were rated as high, moderate, or low risk of bias based on their responses to these questions. A “yes” response indicates a low risk of bias, “no” indicates a high risk of bias, and “unclear” indicates an uncertain risk of bias. Studies with more “yes” responses are rated as having lower risk of bias, while those with more “no” responses are rated as having higher risk of bias. “Unclear” responses indicate that more information is needed to determine the risk of bias in the study. The quality assessment was performed independently by two reviewers, and any disagreements were resolved by consensus or through consultation with a third reviewer. Studies with a high risk of bias were excluded not excluded from the systematic review.

The study selection process was summarized using a PRISMA flow diagram. Quantitative data were presented in tables of individual studies and in summary tables. The quality scores of bias for each eligible study were reported accordingly. We followed the Preferred Reporting Items for Systematic Reviews and Meta-Analyses (PRISMA) guidelines for reporting our systematic review, and our protocol was registered on PROSPERO (CRD42022369954).

## Results

### Included Studies

A total of 63 articles were identified through the initial database search. After removing duplicates, 42 articles remained. After screening the titles and abstracts of these articles, 23 articles were excluded for not meeting the inclusion criteria. The full texts of the remaining 19 articles were assessed for eligibility, and 17 of them were subsequently excluded for not meeting the inclusion criteria. Finally, 2 studies were included in the systematic review. The screening process is summarized in a PRISMA flow diagram (Figure 1). One of the studies was prospective and another retrospective observational study that investigated the incidence and / prevalence of Type 1 diabetes mellitus in children and adolescents in Tanzania. The studies were conducted in Dar es Salaam, Mwanza, Zanzibar, Kilimanjaro Tanzania and were published in 1993 and 2019, respectively.

**Figure 1.**
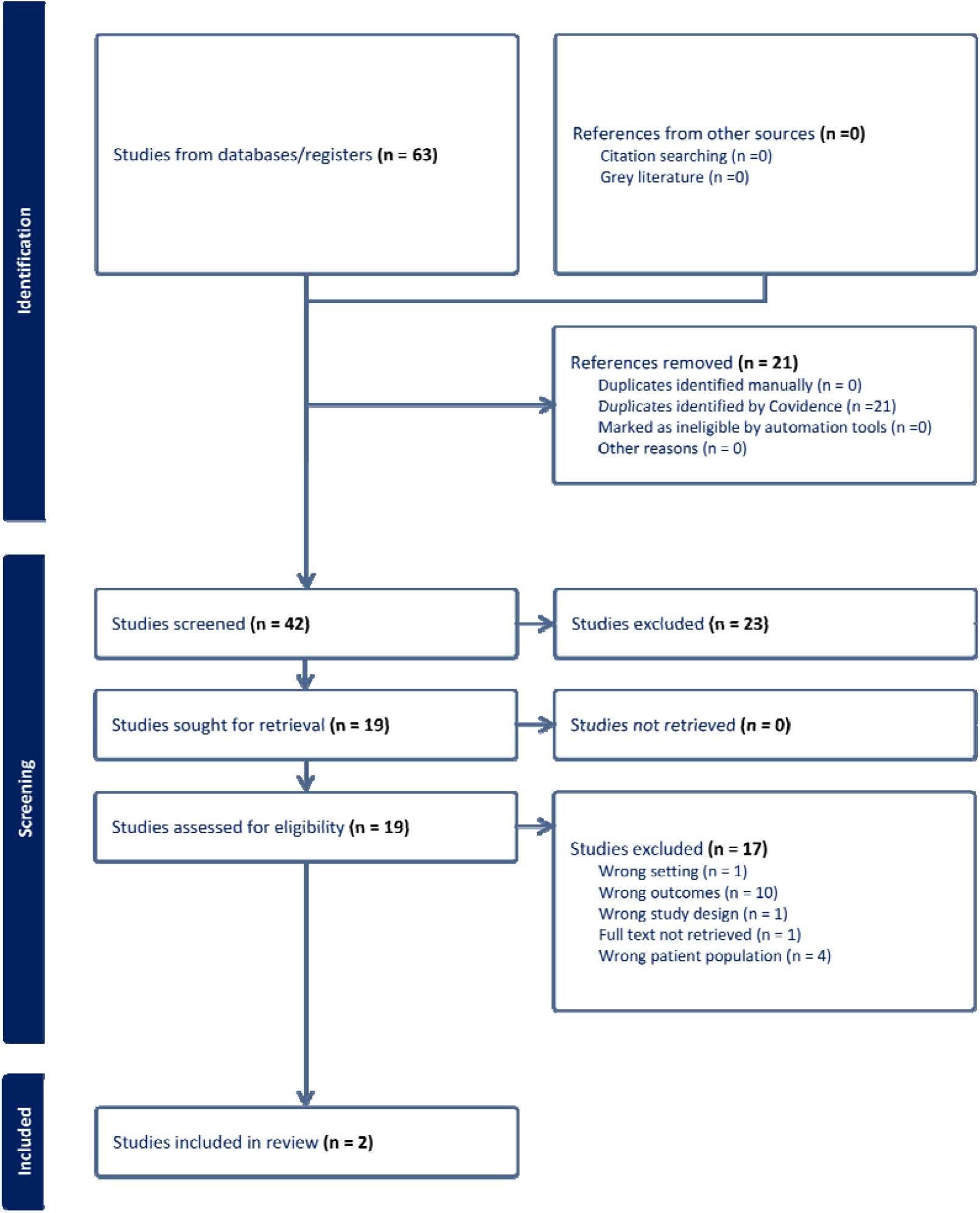
PRISMA flow chart

### Prevalence and Incidence of T1DM in Tanzania

Swai et al. (1993) aimed to determine the annual incidence of insulin-treated diabetes in children and adolescents aged 0-19 years in Dar es Salaam, Tanzania, over a 10-year period from January 1982 to December 1991 (5). The study included all patients aged 19 years and under, who were registered between 1 January 1982 and 31 December 1991 and required insulin from the time of diagnosis. The study found that the annual incidence of T1D for both sexes was 1.5 per 100,000 population aged 0-19 years, with a 95% confidence interval of 1.3 to 1.7 (table 1 below). The study authors noted that T1D is relatively rare in Tanzania and suggested that genetic factors may play a protective role.

**Table 1:**
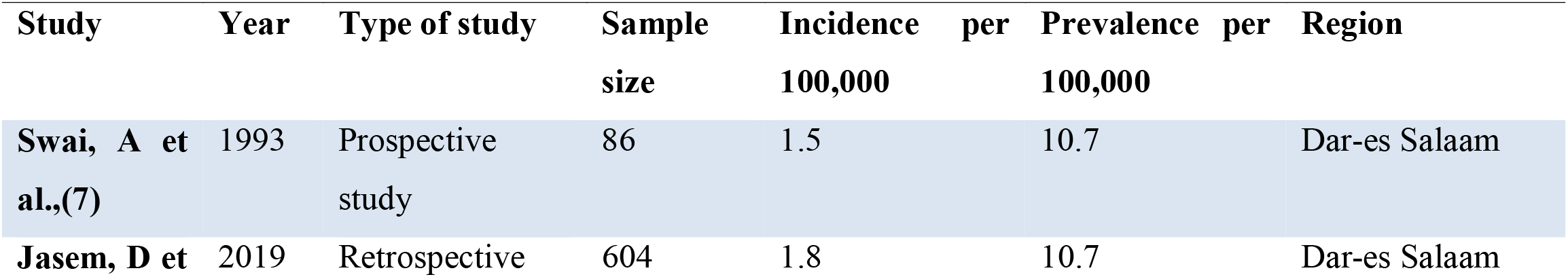
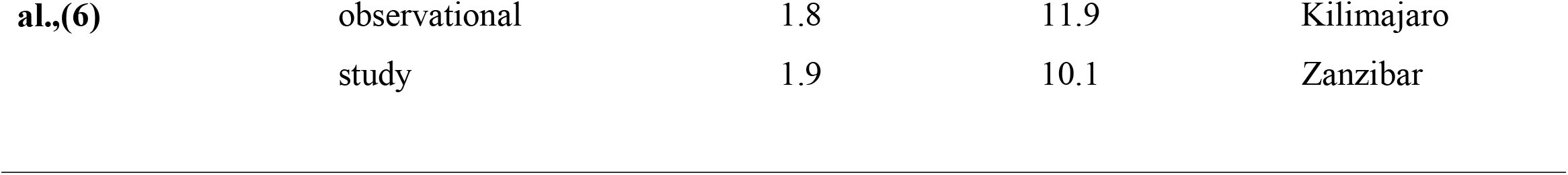
Characteristics of Incidence studies of type 1 diabetes.

**Table 2:** Joanne Briggs critical appraisal tool for risk of bias (Y - Yes, N - No, U – Unclear)

| Authors | Qn 1 | QN 2 | QN 3 | QN 4 | QN 5 | QN 6 | QN 7 | QN 8 | QN 9 | Risk |
| --- | --- | --- | --- | --- | --- | --- | --- | --- | --- | --- |
| Swai et al. (1993) | U | Y | U | N | N | U | U | U | U | Moderate risk |
| Jasem et al. (2019) | U | U | U | Y | N | N | N | N | U | Moderate risk |

The study by Jasem et al. aimed to determine the incidence, prevalence, and clinical manifestations of T1D in Tanzania (6). The study was a retrospective observational study based on medical recordings from January 2010 to April 2016 of diabetic patients attending five hospitals with diabetes services for children in Tanzania. The study included 604 patients’ files, and the overall sex distribution among the patients was similar among females (52%) and males (48%). The prevalence of diabetes among children <15 years old ranged from 10.1 to 11.9 per 100,000 children, and the annual incidence was 1.8-1.9/100,000 children, with peak incidence at 10-14 years. The prevalence was estimated to be 10.7/100,000 children in Dar es Salaam Region (MNH, Temeke), 11.9/100,000 in Kilimanjaro Region (KCMC), and 10.1/100,000 in the Zanzibar Island.

The majority of patients presented with typical signs and symptoms of T1D, and 83.7% of patients had plausible ketoacidosis (DKsA) at diagnosis. Clinical presentation at onset included supposed diabetic keto-acidosis (sDKA) (either measured pH < 7.30 and/or nausea and vomiting, lethargy or drowsiness), polyuria, polydipsia, and weight loss. Overall, the study found that the incidence and prevalence of diabetes in children in Tanzania were relatively low. The authors emphasized the need for increased awareness of diabetes in the healthcare system and community to improve the management and care of diabetes in low resource settings.

### Risk of bias

Based on the Joanne Briggs Institute critical appraisal tool (8), the included studies were assessed for their risk of bias in order to evaluate the quality of evidence presented in this systematic review. No study was excluded based on the quality.

## Discussion

The present systematic review aimed to identify and synthesize the available evidence on the incidence and prevalence of T1D in children and adolescents in Tanzania. The results of the review indicate that only two studies met the inclusion criteria and were included in the analysis.

The first study by Swai et al. (1993) reported an annual incidence of T1D of 1.5 per 100,000 population aged 0-19 years in Dar es Salaam, Tanzania, over a 10-year period from 1982 to 1991. This incidence rate was lower than that reported in developed countries (9–11). The authors suggested that genetic factors may play a protective role, which could contribute to the lower incidence of T1D in Tanzania. The results of the study by Swai et al. are consistent with previous reports of low incidence rates of T1D in sub-Saharan Africa (12). However, it is important to note that the study was conducted in a single urban area and may not be representative of the entire population of Tanzania. The authors’ suggestion of genetic factors playing a protective role should be interpreted with caution as the study did not include a genetic analysis.

The second study by Jasem et al. (2019) reported a prevalence of diabetes among children <15 years old ranging from 10.1 to 11.9 per 100,000 children and an annual incidence of 1.8-1.9/100,000 children in Tanzania. The majority of patients presented with typical signs and symptoms of type 1 diabetes, and 83.7% of patients had plausible ketoacidosis at diagnosis findings that were also comparable to other recently published studies (13–15). The study also found that the prevalence and incidence of diabetes were relatively low in Tanzania.

## Conclusion

In conclusion, the two studies identified in this systematic review provide valuable information on the incidence and prevalence of T1D in Tanzania. Despite the differences in study design and timeframe, both studies suggest that the incidence and prevalence of T1D in this region are relatively low compared to other sub-Saharan countries. However, it is worth noting that these studies are limited, and more research is needed to establish more comprehensive data on the incidence and prevalence of T1D in Tanzania. In particular, future studies should include more regions and utilize standardized diagnostic criteria to enable better comparison of data. The findings of this review highlight the rising need for prevalence and incidence studies. Also, the need for increased awareness of type 1 diabetes in the healthcare system and community to improve diagnosis in low resource settings.

## Data Availability

The datasets used for analysis in this systematic review are available upon reasonable request.

## Limitations

The limitations of this review include the limited number of studies that met the inclusion criteria, which were all conducted in specific geographic locations and may not be representative of the entire population of Tanzania. The studies also had differing methodologies and may not be directly comparable. Additionally, the studies were conducted at different time points, which may not reflect current trends in the prevalence and incidence of T1DM in Tanzania.

## Implication for practice and research

The findings of this systematic review have several implications for clinical and research practice in Tanzania. The limited evidence highlights the need for more prevalence and incidence studies to provide a better understanding of the burden of T1DM in Tanzania. Such studies can inform the development of diabetes management strategies, including screening and early detection programs, to improve the management and control of T1DM in the country.

Given the challenges in resource-limited settings, it is also important to explore cost-effective and culturally appropriate approaches to diabetes screening. Community-based interventions, such as peer support groups and diabetes education programs, have shown promise in improving diabetes awareness in other resource-limited settings and may be worth exploring in Tanzania. Overall, the findings of this systematic review underscore the importance of prioritizing diabetes care and management in Tanzania, and the need for more research to better understand the burden of T1DM in the country.

## Acknowledgement

The authors would like to acknowledge that no funding was received for this study. We would also like to thank the authors of the studies included in this systematic review for their valuable contributions to the field of type 1 diabetes mellitus in Tanzania.

## References

1. Merger SR, Leslie RD, Boehm BO. The broad clinical phenotype of Type 1 diabetes at presentation. Diabet Med. 2013 Feb;30(2):170–8.

2. Karjalainen J, Salmela P, Ilonen J, Surcel HM, Knip M. A comparison of childhood and adult type I diabetes mellitus. N Engl J Med. 1989 Apr 6;320(14):881–6.

3. Ludvigsson J, Edna M, Ramaiya K. Type 1 diabetes in low and middle-income countries -Tanzania a streak of hope. Frontiers in Endocrinology [Internet]. 2023 [cited 2023 Apr 9];14. Available from: https://www.frontiersin.org/articles/10.3389/fendo.2023.1043370

4. Katte JC, McDonald TJ, Sobngwi E, Jones AG. The phenotype of type 1 diabetes in sub-Saharan Africa. Front Public Health. 2023;11:1014626.

5. Swai A.B.M., McLarty D.G., Mugusi F. Tuberculosis in diabetic patients in Tanzania. TROP DOCT. 1990;20(4):147–50.

6. Jasem D, Majaliwa ES, Ramaiya K, Najem S, Swai ABM, Ludvigsson J. Incidence, prevalence and clinical manifestations at onset of juvenile diabetes in Tanzania. Diabetes Research and Clinical Practice [Internet]. 2019 Oct [cited 2023 Apr 7];156:107817. Available from: https://linkinghub.elsevier.com/retrieve/pii/S0168822719308381

7. Swai AB, Lutale JL, McLarty DG. Prospective study of incidence of juvenile diabetes mellitus over 10 years in Dar es Salaam, Tanzania. BMJ [Internet]. 1993 Jun 12 [cited 2023 Apr 7];306(6892):1570–2. Available from: https://www.bmj.com/lookup/doi/10.1136/bmj.306.6892.1570

8. Critical Appraisal Tools | JBI [Internet]. [cited 2023 Apr 9]. Available from: https://jbi.global/critical-appraisal-tools

9. Mobasseri M, Shirmohammadi M, Amiri T, Vahed N, Hosseini Fard H, Ghojazadeh M. Prevalence and incidence of type 1 diabetes in the world: a systematic review and meta-analysis. Health Promot Perspect [Internet]. 2020 Mar 30 [cited 2023 Apr 9];10(2):98–115. Available from: https://www.ncbi.nlm.nih.gov/pmc/articles/PMC7146037/

10. Gomez-Lopera N, Pineda-Trujillo N, Diaz-Valencia PA. Correlating the global increase in type 1 diabetes incidence across age groups with national economic prosperity: A systematic review. World J Diabetes. 2019 Dec 15;10(12):560–80.

11. Green A, Hede SM, Patterson CC, Wild SH, Imperatore G, Roglic G, et al. Type 1 diabetes in 2017: global estimates of incident and prevalent cases in children and adults. Diabetologia. 2021 Dec 1;64(12):2741–50.

12. Hall V, Thomsen RW, Henriksen O, Lohse N. Diabetes in Sub Saharan Africa 1999-2011: Epidemiology and public health implications. a systematic review. BMC Public Health [Internet]. 2011 Jul 14 [cited 2023 Apr 7];11(1):564. Available from: 10.1186/1471-2458-11-564

13. Elendu C, David JA, Udoyen AO, Egbunu EO, Ogbuiyi-Chima IC, Unakalamba LO, et al. Comprehensive review of diabetic ketoacidosis: an update. Ann Med Surg (Lond). 2023 May 23;85(6):2802–7.

14. Gosmanov AR, Gosmanova EO, Kitabchi AE. Hyperglycemic Crises: Diabetic Ketoacidosis and Hyperglycemic Hyperosmolar State. In: Feingold KR, Anawalt B, Blackman MR, Boyce A, Chrousos G, Corpas E, et al., editors. Endotext [Internet]. South Dartmouth (MA): MDText.com, Inc.; 2000 [cited 2024 Feb 22]. Available from: http://www.ncbi.nlm.nih.gov/books/NBK279052/

15. Gallagher E, Siu HYH. Diabetic ketoacidosis as first presentation of type 1 diabetes mellitus in a young child. Can Fam Physician. 2020 Jun;66(6):425–6.

